# Outcome Disparities by Insurance Plan and Educational Attainment in Patients with Atrial Fibrillation

**DOI:** 10.1101/2021.04.25.21256068

**Authors:** Sirin Apiyasawat, Tomon Thongsri, Kulyot Jongpiputvanich, Rungroj Krittayaphong, for the COOL-AF Investigators

## Abstract

**Objectives:** Despite efforts to cut poverty, improve public health, and raise educational attainment, countries still suffer from disparities. Since 2002, Thailand has successfully implemented the Universal Health Coverage Scheme (UCS) to eradicate health care access inequity. Here, we explored the interlink between insurance plan, educational attainment, and adverse clinical outcomes in the national registry of patients with atrial fibrillation (AF) in Thailand.

**Design:** A nationwide prospective multicenter cohort of non-valvular 3402 AF.

**Primary Outcomes:** Patients were followed for 36 months for primary outcomes of all-cause mortality, ischemic stroke, and major bleeding. Survival analysis was performed using restricted mean survival time (RMST) and adjusted with multiple covariates.

**Results:** Data from 3026 AF patients (mean age 67, 59% male) were analyzed. The mean CHA_2_DS_2_VASc and HASBLED scores were 3.0 (SD 1.7) and 1.5 (SD 1.0) respectively. Most of the patients attained the elementary level of education (N=1739, 57.4%). The major health insurance plans were Civil Servant Medical Benefit Scheme (CSMBS; N=1397, 46.2%) and UCS (N=1333, 44.1%). After 36 months of follow-up, 248 patients died (8.2%), 95 suffered from ischemic stroke (3.1%), and 136 suffered from major bleeding (4.5%). AF patients with no formal education lost 1.78 months before they died (adjusted RMST difference -1.78; 95% CI, -3.25 to -0.30; *P* =.02) and 1.04 months before they developed ischemic stroke (adjusted RMST difference -1.04; 95% CI, -2.03 to -0.04; *P* =.04) compared to those with higher education. Educational attainment level was not associated with major bleeding. Across all types of insurance plan, RMSTs to all three clinical outcomes were essentially similar.

**Conclusion:**

1. Education attainment level was independently associated with all-cause mortality and ischemic stroke in AF patients.
2. Health insurance plans were not associated with adverse clinical outcomes.

**Clinical Trial Registration:** Thai Clinical Trial Registration; Study ID: TCTR20160113002

**Strengths and limitations of this study:**

- The study links two of the most important of sociodemographic factors, education and health insurance plan, and clinical outcomes of atrial fibrillation (AF), one of the most common cardiovascular conditions.
- Data is derived from a large prospective and nationwide cohort.
- The study evaluates healthcare quality in universal health coverage system.
- Causal relationship cannot be assumed from a cohort trial.
- Enrollment took place in the medical center, thus, excluding patients who could not access the healthcare facilities.

## Introduction

Poverty, illiteracy, and health adversity feeds each other, and forms a vicious cycle that hinders the progress and prosperity of the nation.^1^ Over the past 2 decades, Thailand has implemented numerous measures to eradicate these undesirable states. ^2-4^ One of the most audacious and yet highly successful policies was the pro-poor medical plan named universal coverage scheme (UCS).^2,5^ The scheme provided a non-contributory benefit for all uninsured citizen.^2^ Since its implementation in 2002, UCS substantially reduced uninsured population from approximately 47 million (30% of total population) to 1.1 million (1.65% of total population) in 2011, and to less than 400,000 people (0.06% of total population) in 2018.^2,6^ UCS was often compared to civil servant medical benefit scheme (CSMBS) for quality and accessibility of care. Offered as a fringe benefit for government officials, CSMBS was regarded as one of the most comprehensive health insurance plans in the country.^2^ CSMBS beneficiaries reportedly had better clinical outcomes than those of UCS.^7-9^ However, such gaps were closing while UCS continued to strengthen its primary health care system and expand its benefit packages.^2^

Along with healthcare, education is another extremely vital tool to break the vicious cycle.^1,10^ Education level and literacy rate was one of the major determinants of all-cause mortality, cardiovascular mortality, and cancer mortality.^11,12^ Low level of education limited understanding of one’s own illness.^13^ In atrial fibrillation (AF), such that limitation led to poor medication adherence,^14^ worsened symptom severity,^15^ and high AF related complications.^14,16^ Here, we explored the interlink between insurance plan, educational attainment, and adverse clinical outcomes in the national registry of patients with AF. We believed that with the success of UCS, the insurance plan would not be linked to adverse clinical outcomes. Educational attainment, rather, was hypothesized to be associated with risks of mortality, ischemic stroke, and major bleeding.

## Methods

Between 2014-2017, patients with AF across Thailand were consecutively enrolled in the COOL-AF (<u>**Co**</u>hort of Antithrombotic Use and <u>**O**</u>ptimal INR <u>**L**</u>evel in Patients with Non-Valvular <u>**A**</u>trial <u>**F**</u>ibrillation in Thailand) registry. The detail of the study was stated elsewhere.^17^ Briefly, patients age >18 year with electrocardiography confirmed AF were eligible for the enrolment. The exclusion criteria^17^ were (1) ischemic stroke within 3 months; (2) hematologic disorders that can increase the risk of bleeding, such as thrombocytopenia (<100000/mm3) and myeloproliferative disorders; (3) mechanical prosthetic valve or valve repair; (4) rheumatic valve disease or severe valve disease; (5) AF associated with transient reversible cause; (6) current participation in a clinical trial; (7) life expectancy <3 years; (8) pregnancy; (9) inability to attend follow-up visits; and (10) refusal to participate in the study. The protocol was approved by the ethics committee of each participating hospital. All patients provided written informed consent.

### Data Collection

Baseline data including demographics, educational attainment, insurance plan, medical history, and medications taken were obtained at the enrollment from medical records and patient interviews. Patients with missing information on educational attainment or insurance plan were excluded. All patients were prospectively followed for major adverse outcomes of death from any causes, ischemic stroke, and major bleeding. The follow-up data were collected from medical records and telephone interviews every 6 months until the end of follow-up at 36 months.

### Data Classifications

The levels of the educational attainment were elementary, secondary, higher education, and no formal education. The categories of the insurance plans were (1) universal coverage scheme (UCS), a non-contributory insurance scheme provided for all uninsured citizens of Thailand; (2) compulsory social security scheme (SSS), a contributory and compulsory scheme for non-government employees; (3) civil servant medical benefit scheme (CSBMS), a fringe benefit provided for government employees; and (4) non-government-based scheme (NGS) which included private insurance, opt-out of the plan, and out-of-pocket payment. The stroke and bleeding risks were assessed by CHA_2_DS_2_VASc and HASBLED scores respectively according to the standard guidelines.^18^ Types of anticoagulants were grouped into none, vitamin K antagonist (VKA), and direct oral anticoagulant (DOAC).

### Statistical Analysis

All analyses were generated using SAS software, version 9.4_M6 of the SAS OnDemand for Academics (SAS Institute Inc., Cary, NC, USA.). Variables were expressed in number and percentage for categorical type and mean and SD for continuous type. Chi-square and one-way ANOVA was used to compare the differences in baseline characteristics. Event probabilities were assessed by restricted mean survival time (RMST) analysis using LIFETEST and RMSTREG procedures in the SAS/STAT software. RMST is equivalent to an area under the whole survival curve up to a fixed time point. It is widely advocated as an alternative to Cox proportional hazard model especially when proportionality is violated.^19,20^ Here, we computed RMST to the time horizon of 36 months using Kaplan-Meier method for survival probability estimation. We then adjusted RMST with multiple covariates using pseudo-value linear regression model.^21^ The model consisted of age, sex, educational attainment, insurance plan, types of anticoagulant, CHA_2_DS_2_VASc, and HASBLED scores. The results were presented as the differences of RMSTs and 95% confidence interval (CI) for each covariate. The RMST difference estimated months gain or loss of event-free time over a period of 36 months. *P* values were reported for all between-group comparisons. A value < .05 was considered significant.

## Results

The COOL-AF registry enrolled a total of 3402 patients. Of these, 376 patients had incomplete data and were excluded from the study. Thus, 3026 patients remained for the analysis. The population <u>(Table 1)</u> was approximately 67 years of age and primarily male (N=1779, 58.8%). The most common comorbidities were hypertension (N=2061, 68.1%), followed by congestive heart failure (CHF; N=829, 27.4%), and diabetes (N=730, 24.1%). The mean CHA_2_DS_2_VASc and HASBLED scores were 3.0 (SD 1.7) and 1.5 (SD 1.0) respectively.

**Table 1.**
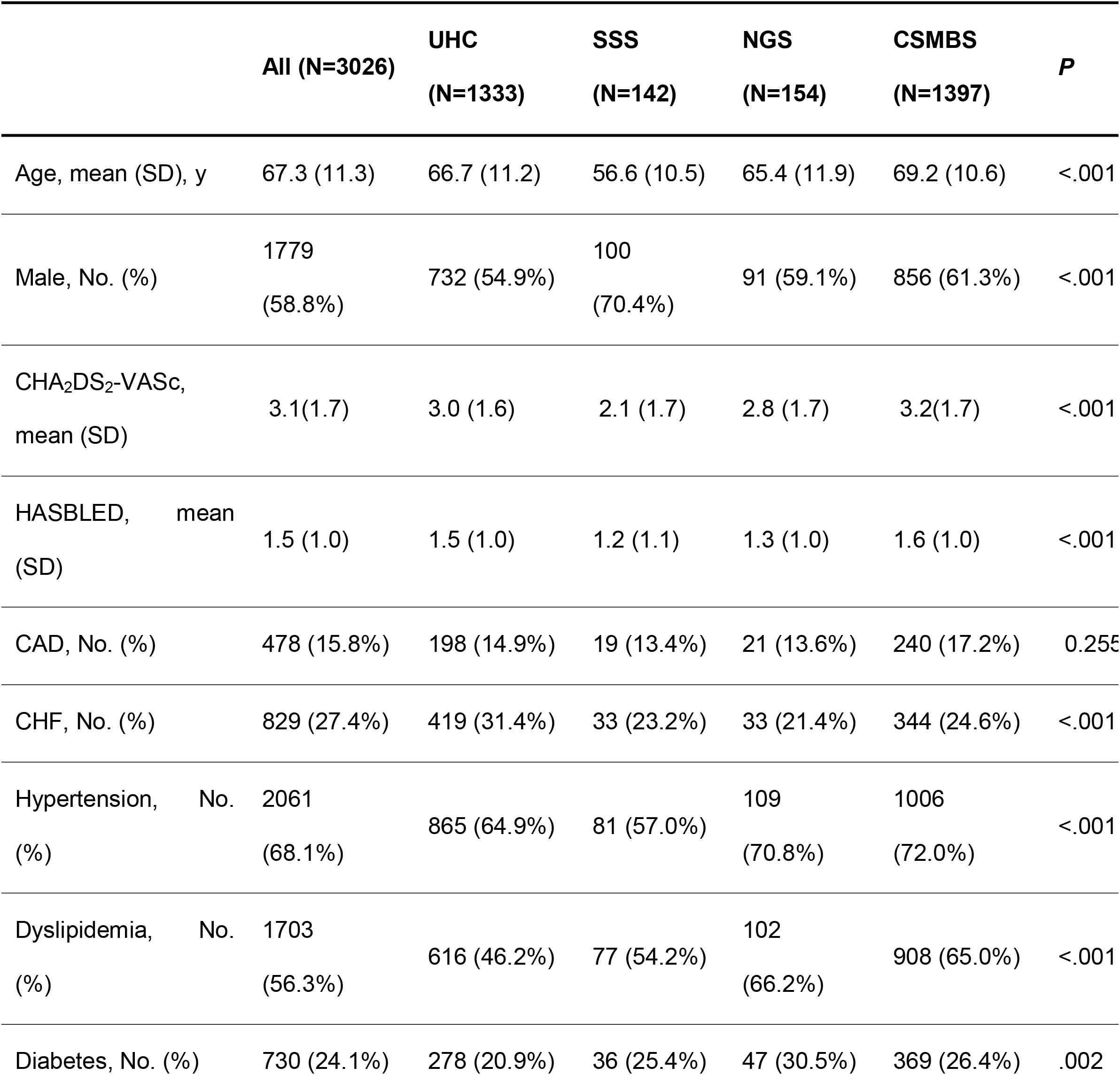
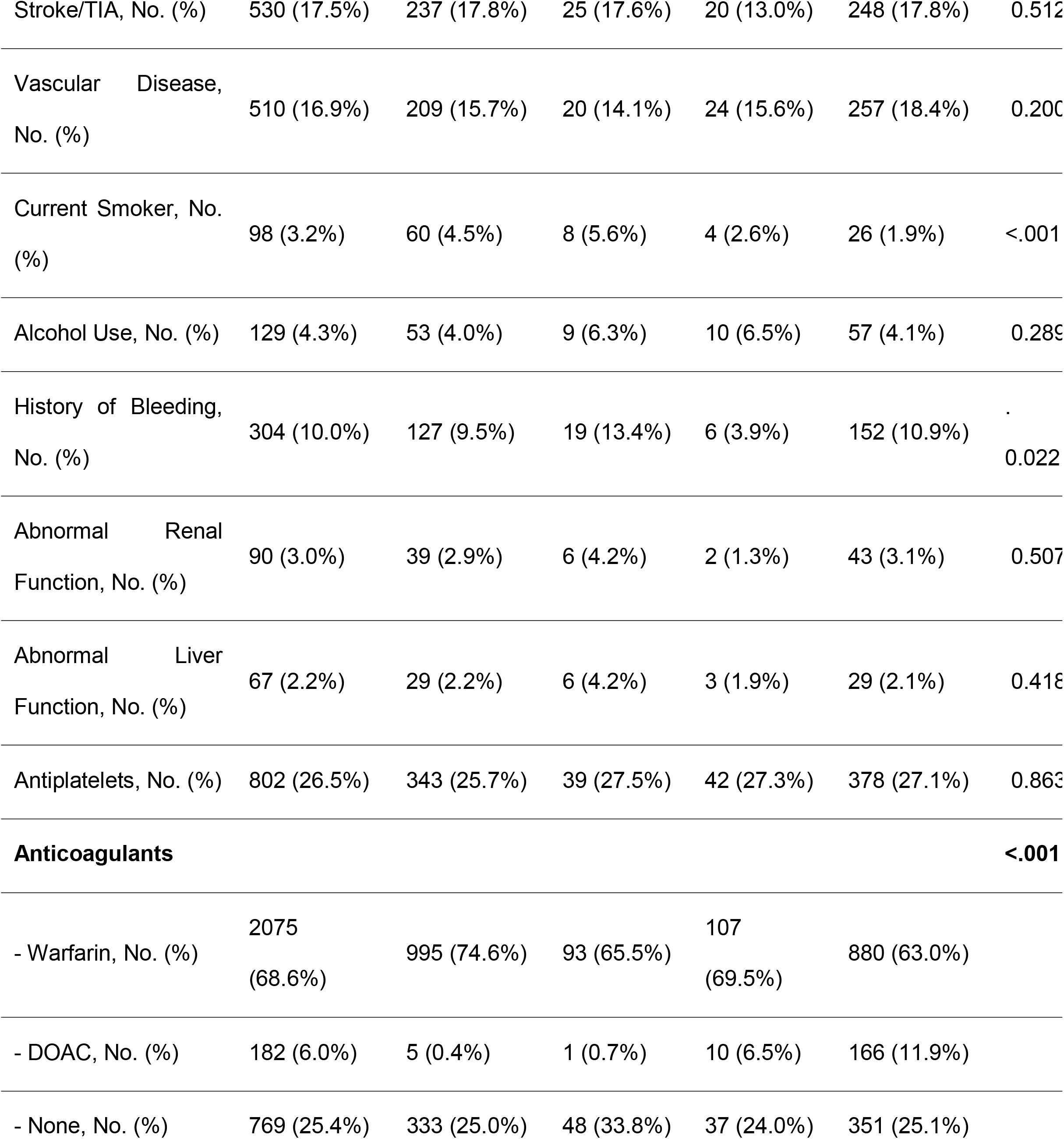
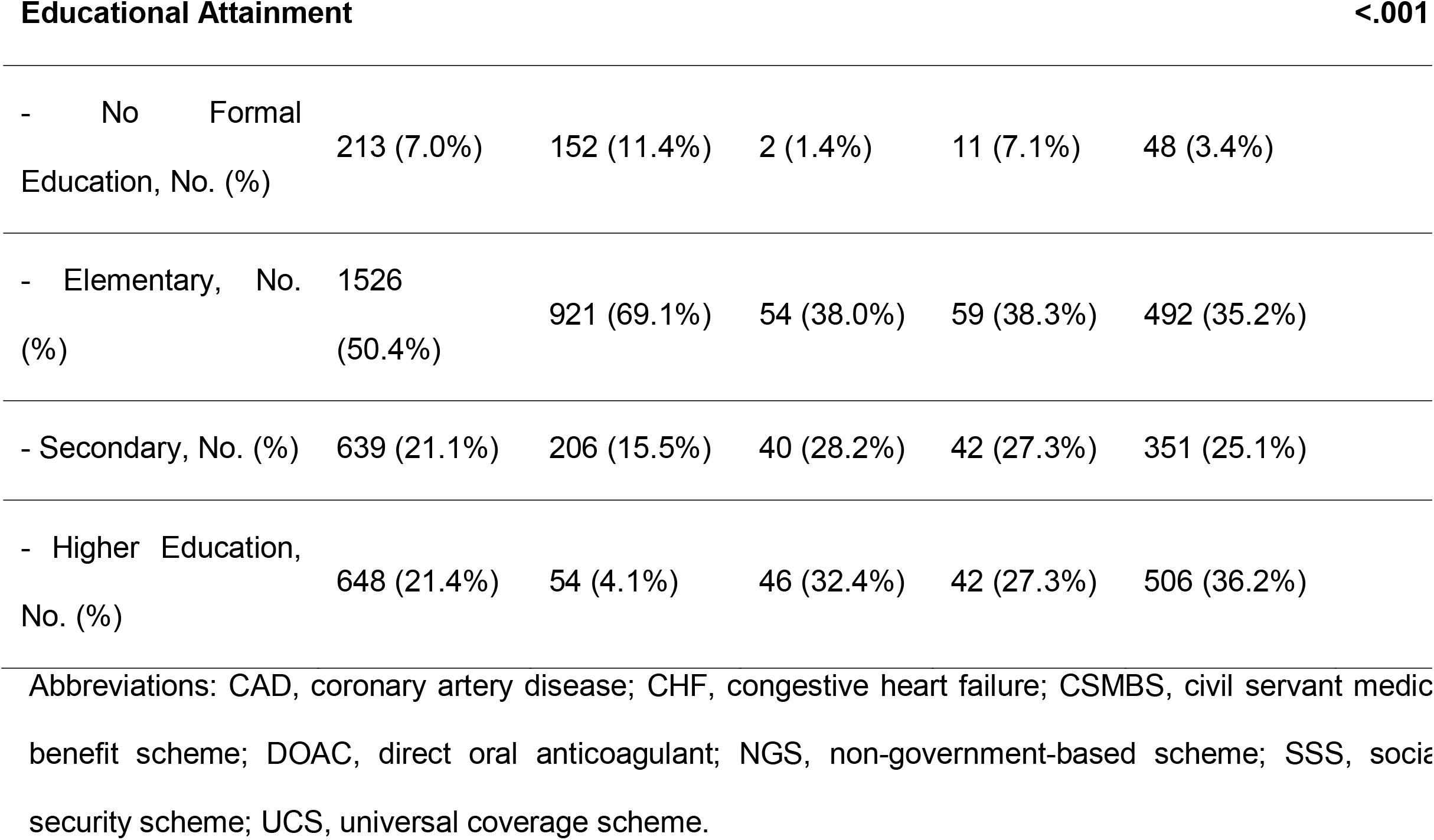
Baseline Characteristics by Insurance Plans (N=3026)

### Insurance Plan (Table 1)

The population was predominantly insured by either CSMBS (N=1397, 46.2%) or UCS (N=1333, 44.1%). Patients insured by CSMBS had higher stroke and bleeding risks assessed by CHA_2_DS_2_VASc (mean 3.2 for CSMBS, 3.0 for UCS, 2.8 for NGS, 2.1 for SSS; *P*<.001) and HASBLED (mean 1.6 for CSMBS, 1.5 for UCS, 1.3 for NGS, 1.2 for SSS; *P*<.001) scores. Approximately 12% of CSMBS insurers were taking DOACs compared to 0.4%, 0.7%, and 6.5% in those insured by UCS, SSS, and NGS respectively (*P*<.001). Higher education was attained more frequently in 36.2% of CSMBS insurers, compared to 32.4% in SSS, 27.3% in NGS, and 4.1% in UCS (*P*<.001).

### Educational Attainment

There was more than one-third of the patients attained at least secondary school (N=1287, 42.5%). Among those, male predominated (N=948, 73.7%). Age, CHA_2_DS_2_VASc and HASBLED scores <u>(Table 2)</u> were at the highest in those with no formal education (mean age 74, CHA_2_DS_2_VASc 3.9, HASBLED 1.7) followed by those with elementary (mean age 69, CHA_2_DS_2_VASc 3.3, HASBLED 1.6), secondary (mean age 64, CHA_2_DS_2_VASc 2.7, HASBLED 1.5), and higher education (mean age 63, CHA_2_DS_2_VASc 2.6, HASBLED 1.4; *P*<.001 for all comparisons). The anticoagulation rate was higher in no formal education group (N=182, 85.4%) than elementary (N=1129, 74.0%), secondary (N=477, 74.6%), and higher education (N=469, 72.4%; *P*<.001).

**Table 2.**
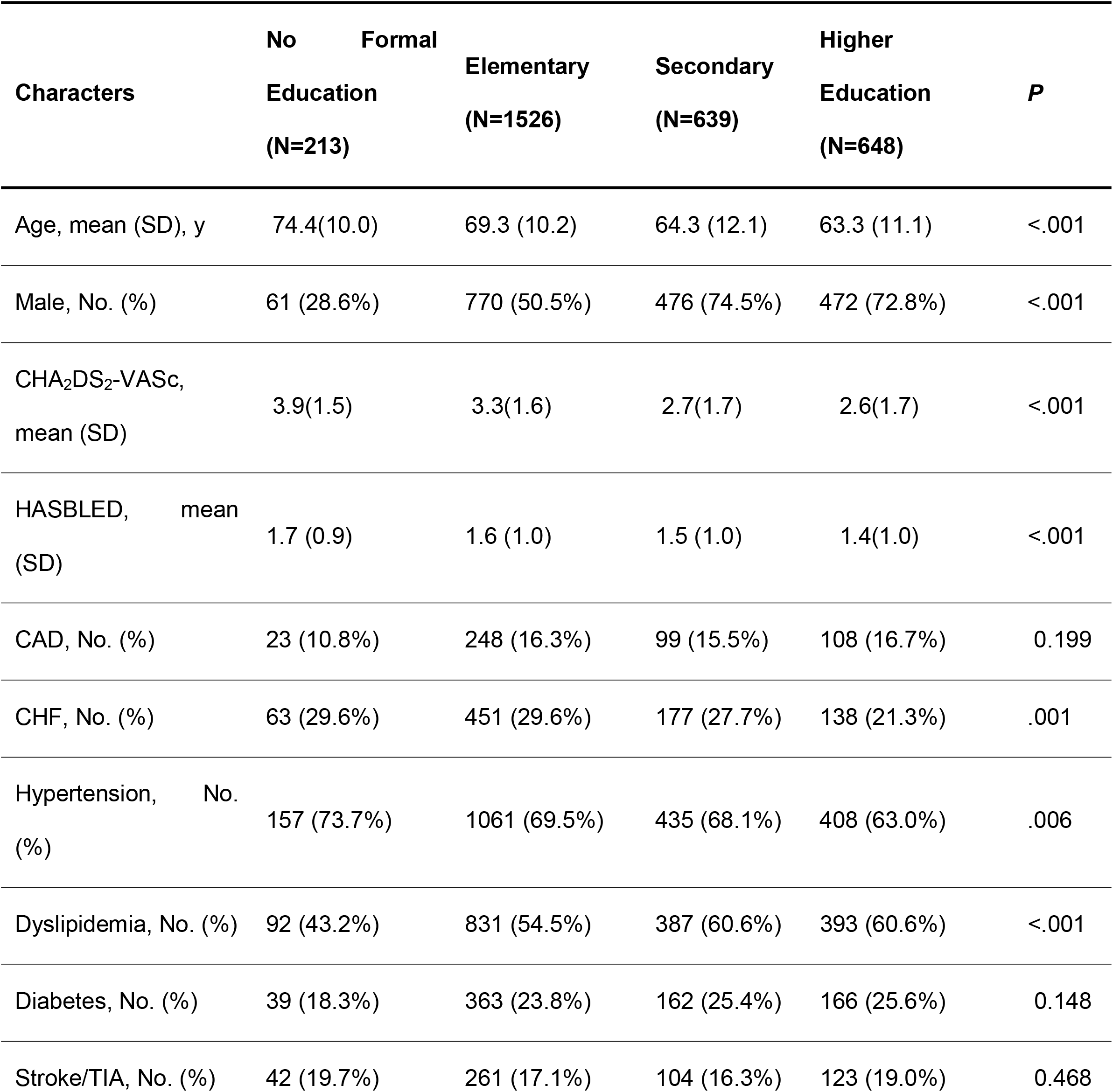
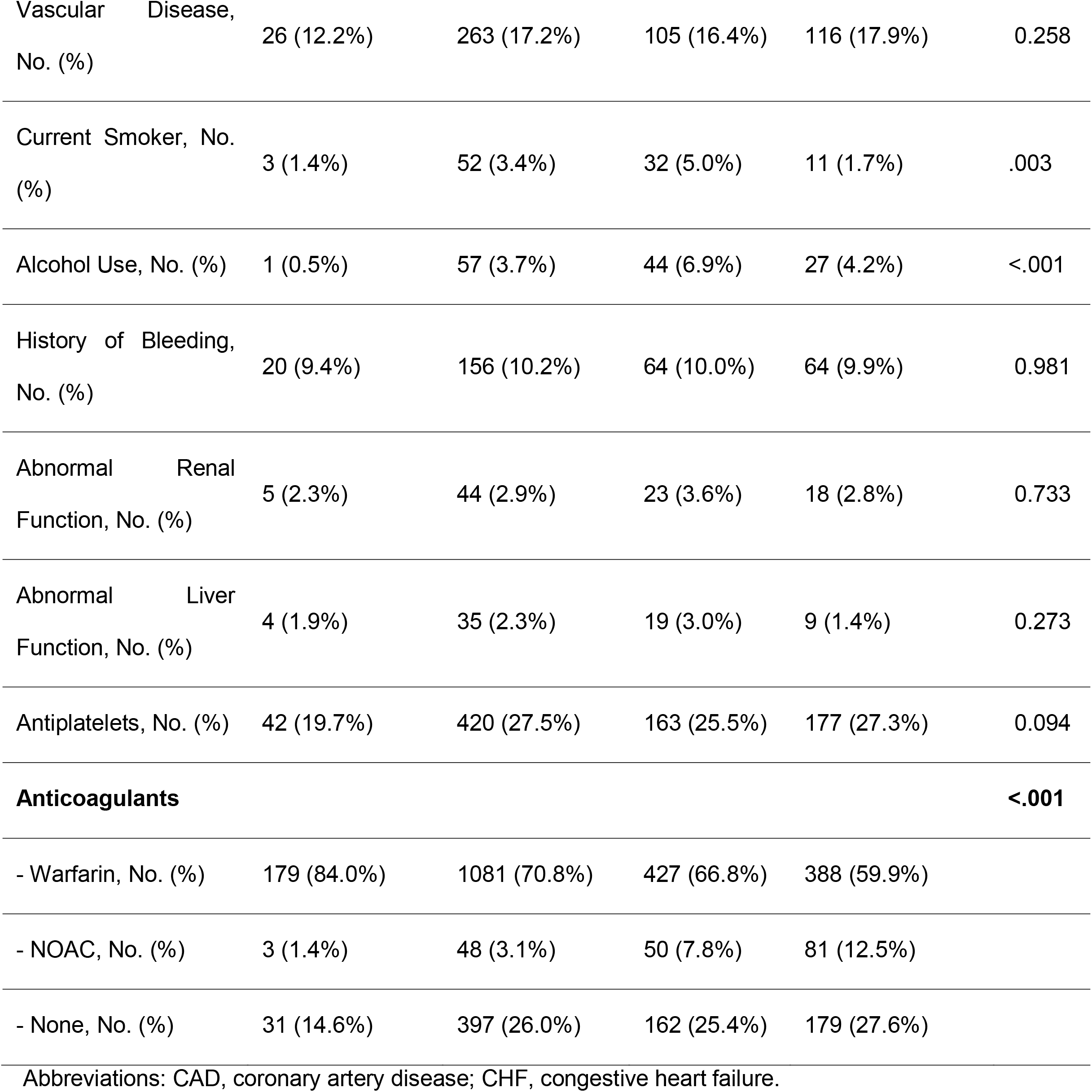
Baseline Characteristics by Educational Attainment (N=3026)

Patients attained secondary school were likely to smoke (5% in secondary, 3.4% in elementary, 1.7% in higher education, 1.4% in no education) and use alcohol (6.9% in secondary, 4.2% in higher education, 3.7% in elementary, 0.5% in no education) than those with other educational attainment (*P*<.001 for both comparisons).

### Adverse Outcomes

After 36 months of follow-up, 248 patients died (8.2%), 95 suffered from ischemic stroke (3.1%), and 136 suffered from major bleeding (4.5%). Figure 1 and figure 2 shows the Kaplan-Meier survival curves of all-cause mortality by insurance plan and educational attainment respectively. Patients with higher educational attainment survived longer than those with lower educational attainment (RMST for higher education vs. secondary vs. elementary vs. no formal education, 35.02 vs. 34.29 vs. 33.70 vs. 31.98 months; *P*<.001). They also experienced longer event-free time to ischemic stroke (RMST for higher education vs. secondary vs. elementary vs. no formal education, 35.58 vs. 35.46 vs. 35.11 vs. 34.15 months; *P*=.004) and major bleeding (RMST for higher education vs. secondary vs. elementary vs. no formal education, 35.31 vs. 34.72 vs. 37.82 vs. 34.18 months; *P*=.03) than those with lower educational attainment (Table 4). RMSTs of all 3 adverse outcomes were not different among plans of insurance (Table 3).

**Table 3.**
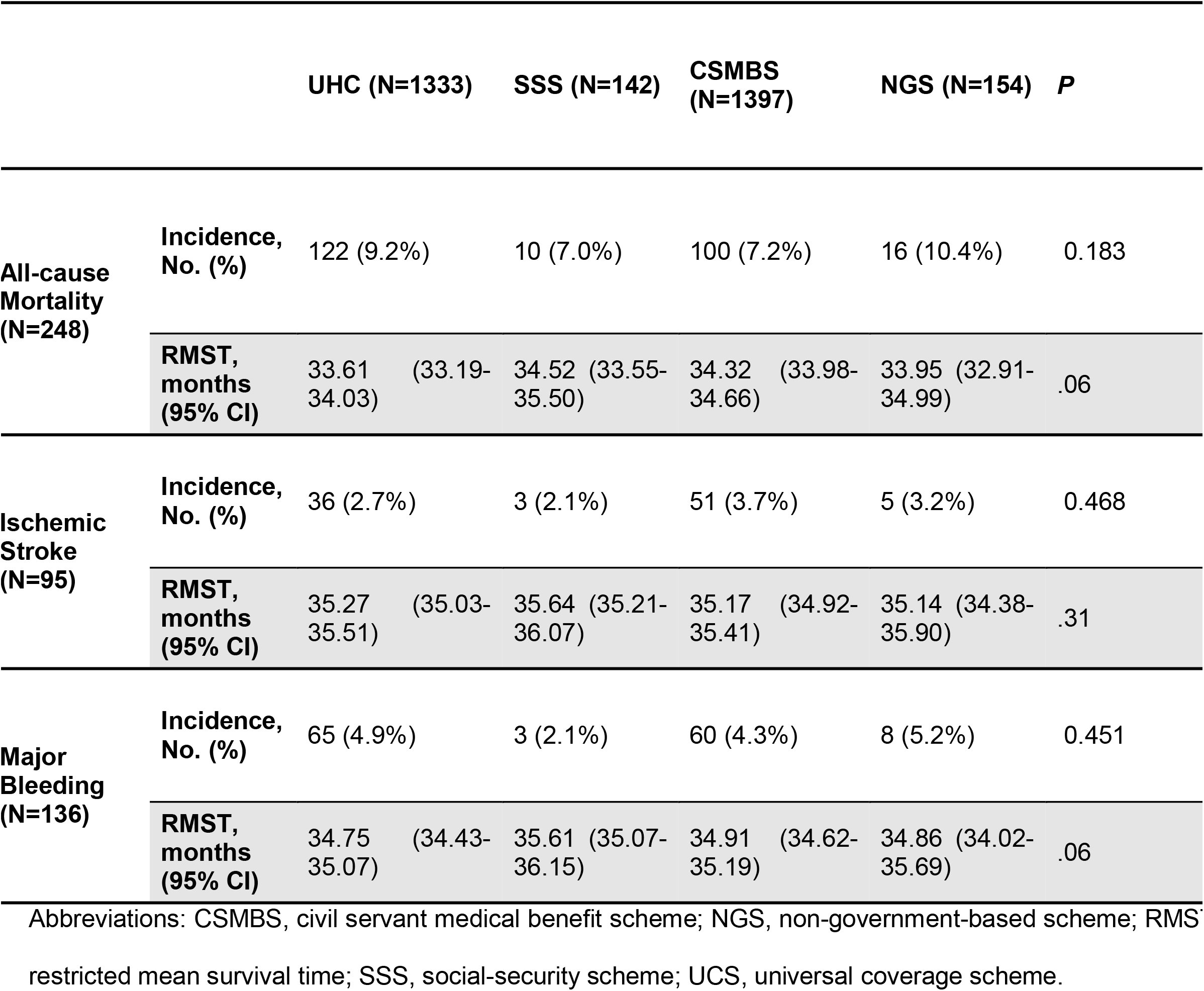
Incidence and Restricted Mean Survival Time of Clinical Outcomes by Insurance Plan.

**Table 4.** Incidence and Restricted Mean Survival Time of Clinical Outcomes by Educational Attainment.

|  |  | No Formal<br>Education<br>(N=213) | Elementary<br>(N=1526) | Secondary<br>(N=639) | Higher<br>Education<br>(N=648) | *P* |
| --- | --- | --- | --- | --- | --- | --- |
| **All-cause**<br>**Mortality**<br>**(N=248)** | **Incidence,**<br>**No. (%)** | 30 (14.1%) | 146 (9.6%) | 45 (7.0%) | 27 (4.2%) | <.001 |
|  | **RMST,**<br>**months**<br>**(95% CI)** | 31.98 (30.62-33.33) | 33.70 (33.32-34.80) | 34.29 (33.78-34.80) | 35.02 (34.65-35.40) | <.001 |
| **Ischemic**<br>**Stroke**<br>**(N=95)** | **Incidence,**<br>**No. (%)** | 14 (6.6%) | 55 (3.6%) | 14 (2.2%) | 12 (1.9%) | .002 |
|  | **RMST,**<br>**months**<br>**(95% CI)** | 34.15 (33.21-35.10) | 35.11 (34.87-35.36) | 35.46 (35.16-35.75) | 35.58 (35.32-35.84) | .004 |
| **Major**<br>**Bleeding**<br>**(N=136)** | **Incidence,**<br>**No. (%)** | 13 (6.1%) | 70 (4.6%) | 35 (5.5%) | 18 (2.8%) | 0.066 |
|  | **RMST,**<br>**months**<br>**(95% CI)** | 34.18 (33.22-35.14) | 34.82 (34.54-35.11) | 34.72 (34.27-35.17) | 35.31 (34.98-35.64) | .03 |

Abbreviations: RMST, restricted mean survival time.

**Figure 1.**
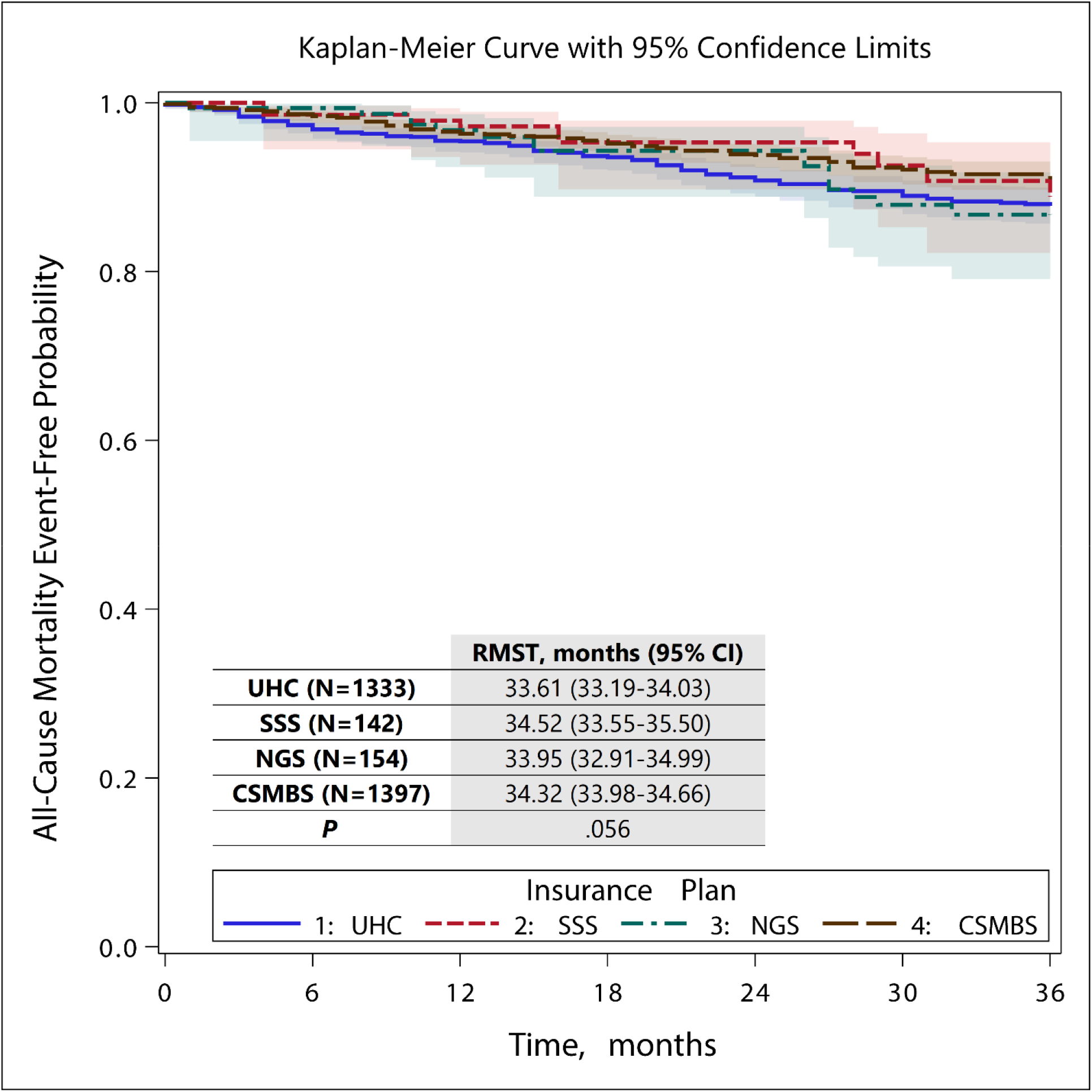
Kaplan-Meier Curve for All-cause Mortality by Insurance Plans Abbreviations: CSMBS, civil servant medical benefit scheme; NGS, non-government-based scheme; SSS, social-security scheme; UCS, universal coverage scheme.

**Figure 2.**
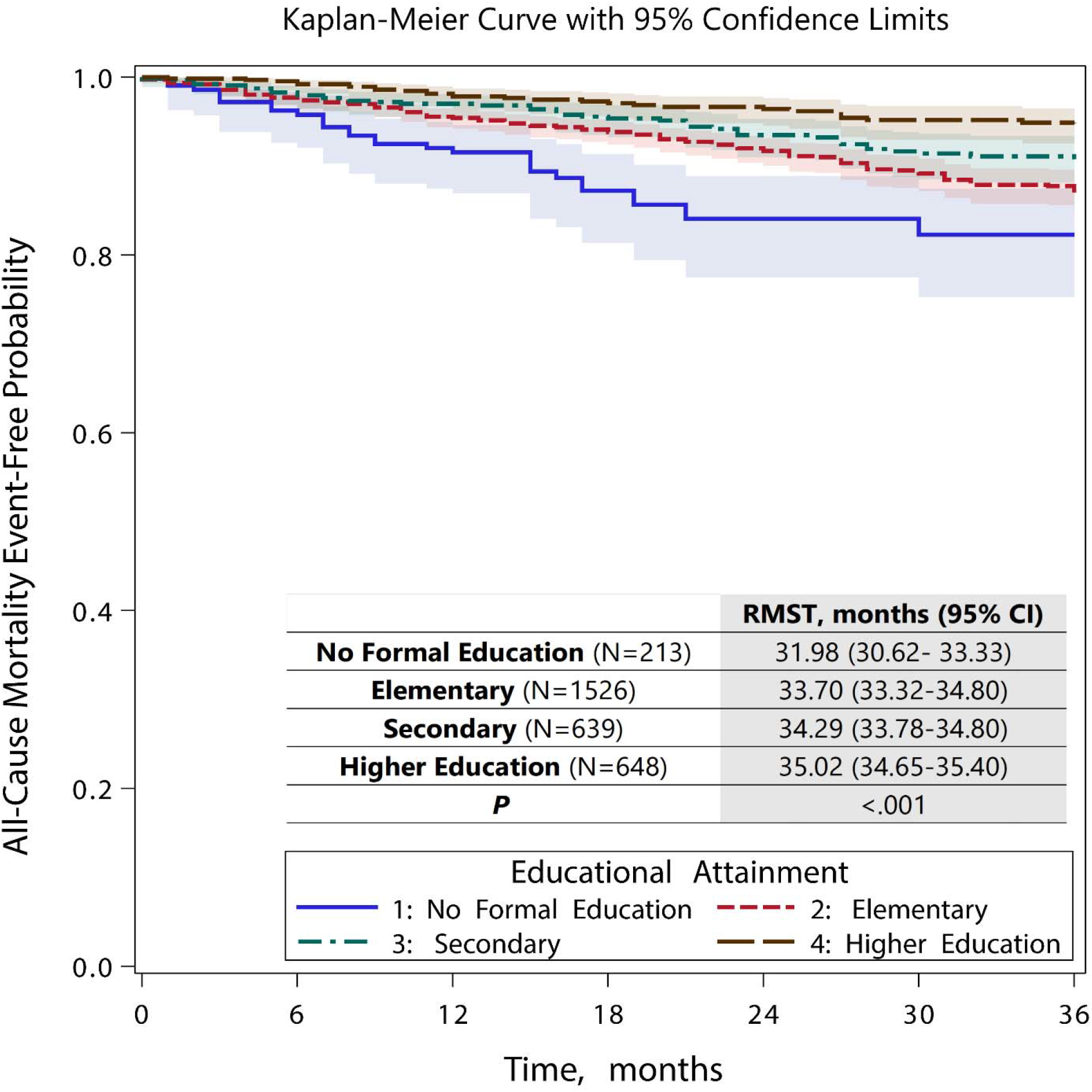
Kaplan-Meier Curve for All-cause Mortality by Educational Attainment

In the multivariate analyses, educational attainment was associated with all-cause mortality and ischemic stroke independent of comorbidities (Figures 3 and 4). Over a period of 36 months, AF patients with no formal education lost 1.78 months before they died (RMST difference −1.78; 95% CI, −3.25 to −0.30; *P*=.02) and 1.04 months before they developed ischemic stroke (RMST difference −1.04; 95% CI, −2.03 to −0.04; *P* =.04) compared to those with higher education. Older age, higher CHA_2_DS_2_VASc, and HASBLED scores also shortened survival time to all-cause mortality. Survival time was lost by 0.45 months for every 10-year increase in age (RMST difference −0.45; 95% CI, −0.8 to −0.1; *P* =.01), 0.35 months for every point increase in CHA_2_DS_2_VASc score (RMST difference −0.35; 95% CI, −0.6 to −0.1; *P* =.01), and 0.41 months for every point increase in HASBLED score (RMST difference −0.41; 95% CI, −0.76 to −0.07; *P* =.02). Use of anticoagulants extended event-free time to ischemic stroke by 0.99 months for DOAC (95% CI, 0.45 to 1.54; *P* = <.001) and 0.69 months for VKA (95% CI, 0.24 to 1.13; *P* = .003). Time to major bleeding (Figure 5) was lost in patients with older age (RMST difference per 10 years −0.35; 95% CI, −0.59 to - 0.11; *P* =.003), male gender (RMST difference −0.68; 95% CI, −1.11 to –0.25; *P* =.002), and use of VKA (RMST difference −0.65; 95% CI, −1.04 to −0.26; *P* = .001). Across all 3 major outcomes, insurance plan was not associated with gain or loss in survival times (Figure 3-5).

**Figure 3.**
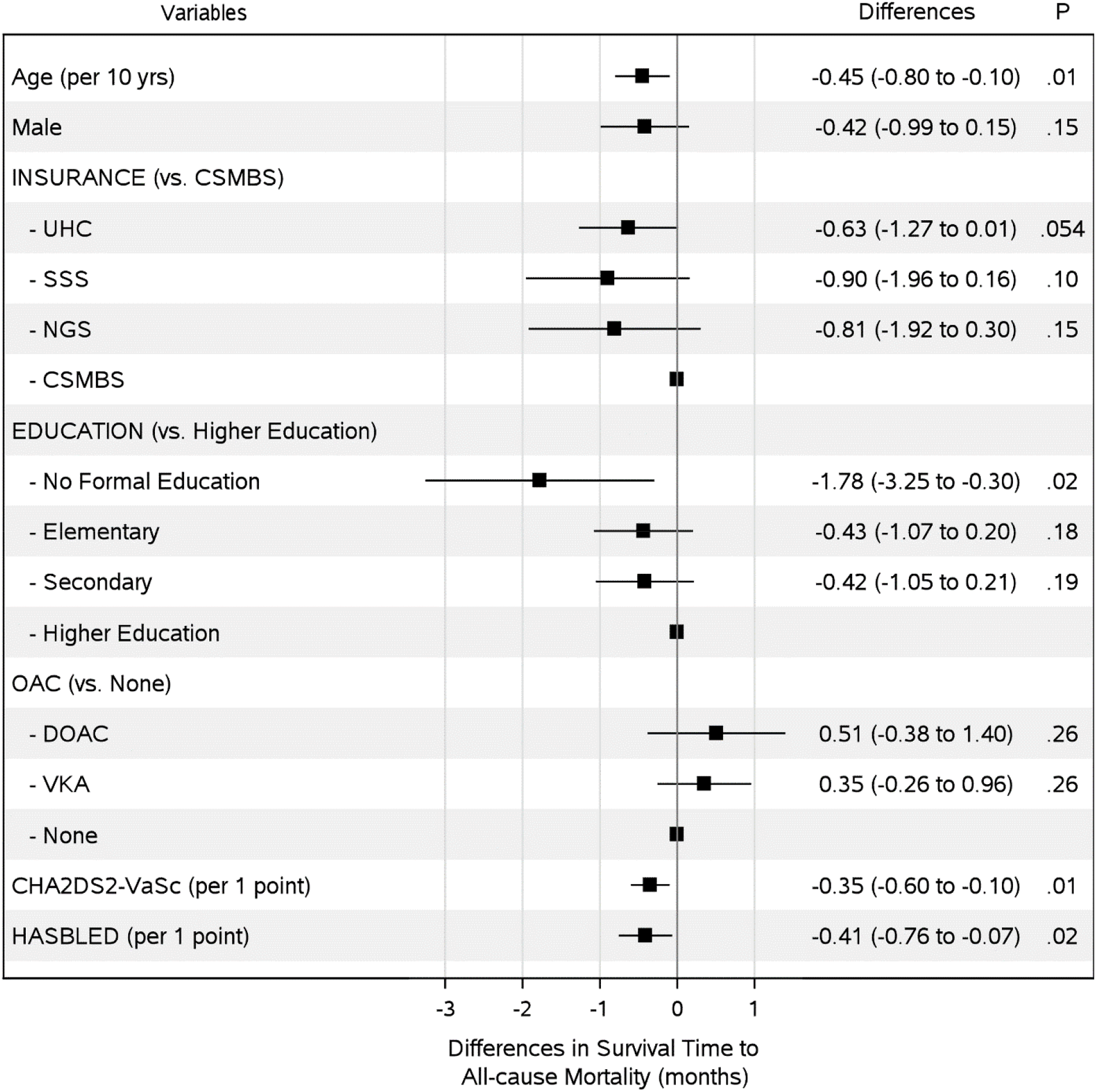
Differences in Adjusted Restricted Median Survival Time to All-Cause Mortality over a Period of 36 Months. Abbreviations: CAD, coronary artery disease; CHF, congestive heart failure; CSMBS, civil servant medical benefit scheme; DOAC, direct oral anticoagulant; NGS, non-government-based scheme; SSS, social-security scheme; UCS, universal coverage scheme; VKA, vitamin K antagonist.

**Figure 4.**
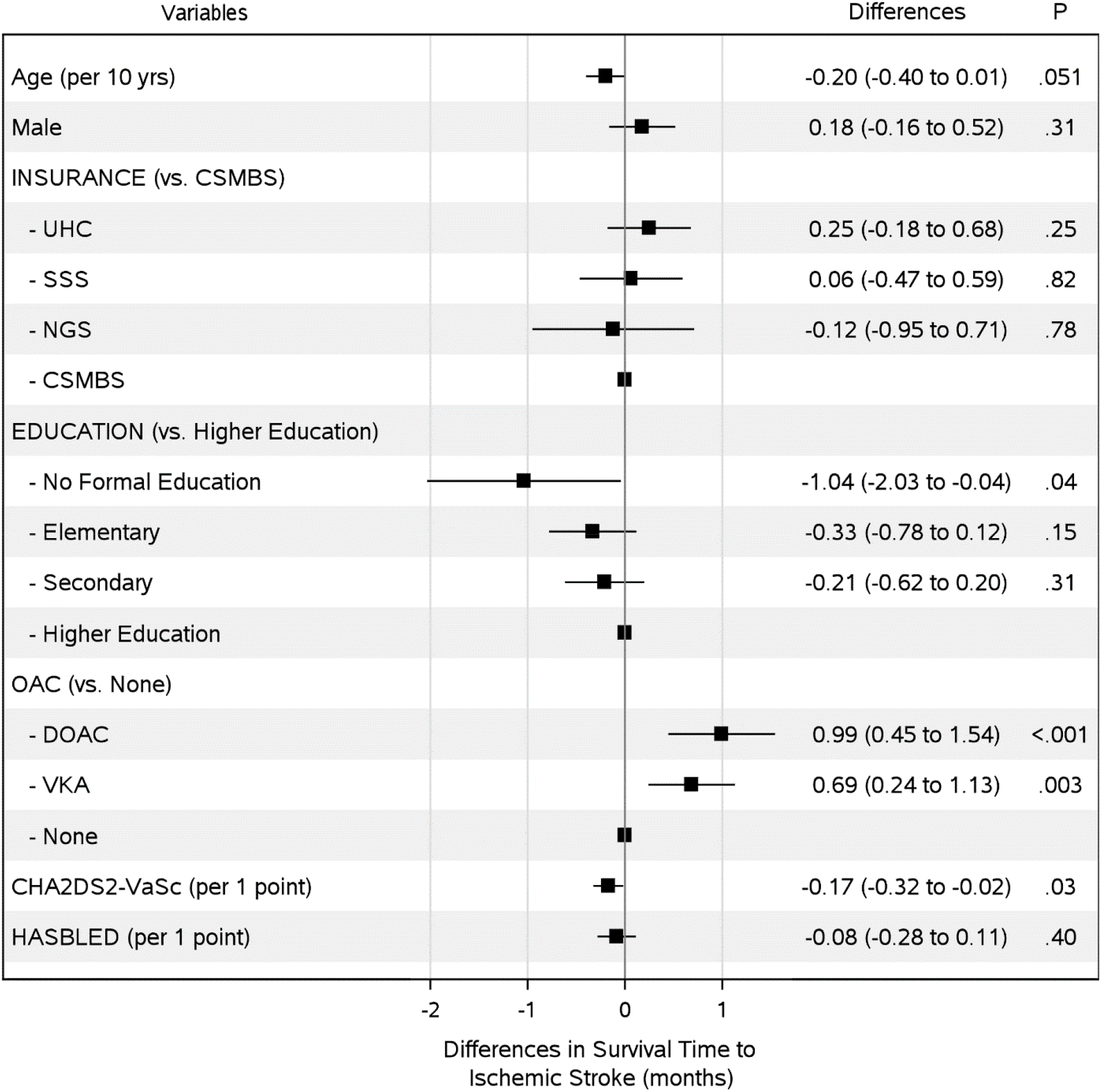
Differences in Adjusted Restricted Median Survival Time to Ischemic Stroke over a Period of 36 Months. Abbreviations: CAD, coronary artery disease; CHF, congestive heart failure; CSMBS, civil servant medical benefit scheme; DOAC, direct oral anticoagulant; NGS, non-government-based scheme; SSS, social-security scheme; UCS, universal coverage scheme; VKA, vitamin K antagonist.

**Figure 5.**
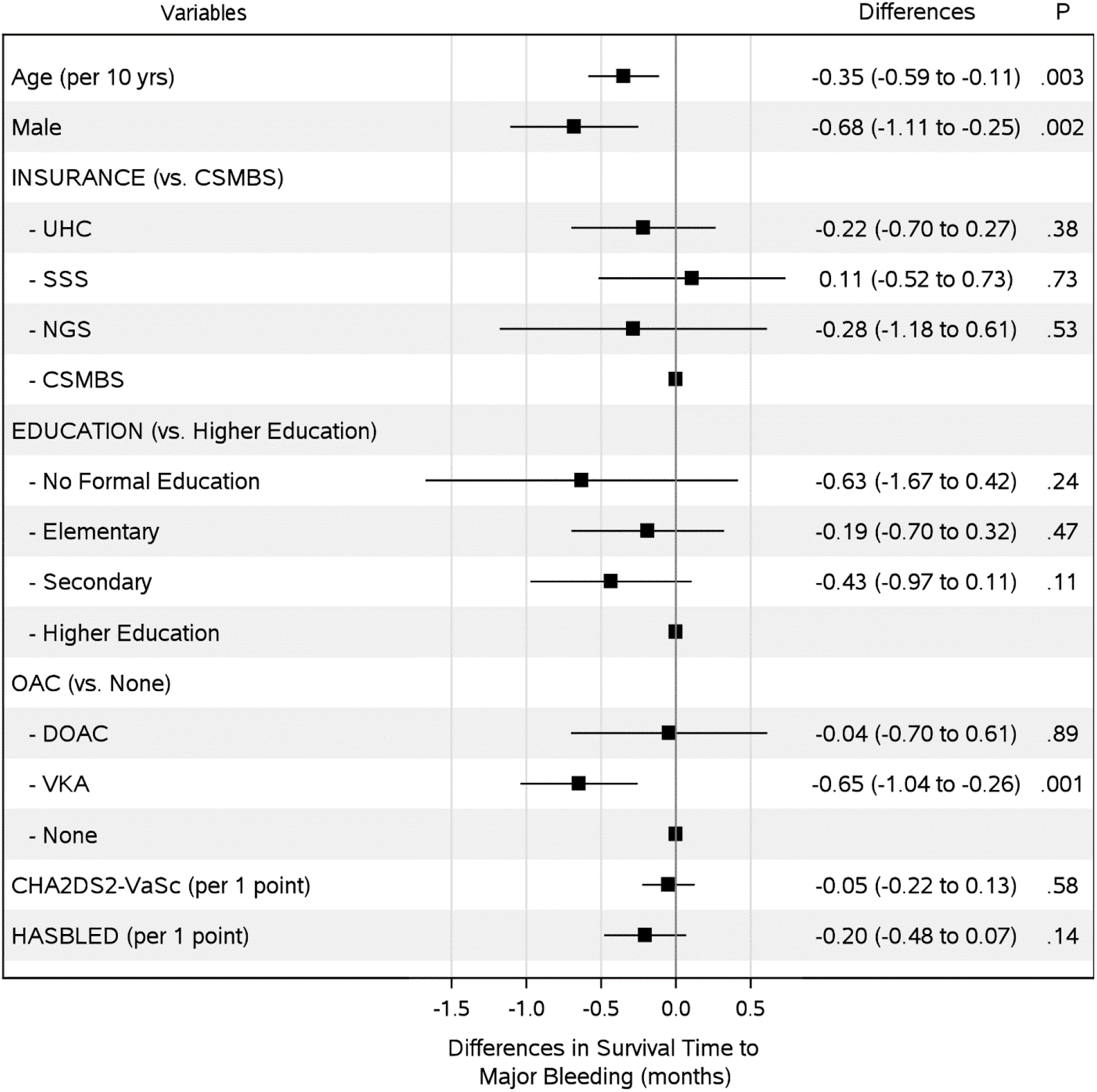
Differences in Adjusted Restricted Median Survival Time to Major Bleeding over a Period of 36 Months. Abbreviations: CAD, coronary artery disease; CHF, congestive heart failure; CSMBS, civil servant medical benefit scheme; DOAC, direct oral anticoagulant; NGS, non-government-based scheme; SSS, social-security scheme; UCS, universal coverage scheme; VKA, vitamin K antagonist.

## Discussion

In the large registry of AF patients across Thailand, educational attainment was associated with all-cause mortality and ischemic stroke independent of multiple covariates. Over 36 months of follow-up, those with no formal education died 1.8 months sooner and suffered stroke 1 month sooner than those with higher education. Types of Insurance plan were not linked to either all-cause mortality, ischemic stroke, or major bleeding.

The significant impact of education on cardiovascular outcome has been shown globally.^16,23-25^ Among Asian population, data from the Asia Pacific Cohort Studies Collaboration^25^ reported that Asians with the lowest educational attainment had a 54% higher risk of stroke and 78% higher risk of cardiovascular mortality than those with tertiary level. Among patients with AF, a large-scale study in Norway reported an educational gradient in mortality between higher education and high school or less.^26^ For other AF outcomes such as stroke or major bleeding, the data on educational disparities was limited. In a patient survey,^27^ higher level of education was associated with better understanding of anticoagulant therapy and lower incidence of bleeding. Here, in a prospective cohort, we reported educational disparities in all-cause mortality and stroke but not major bleeding. The disparities were detected between no formal education and higher education levels independent of age, sex, bleeding risk, stroke risk, insurance plan, and anticoagulant status. Patients with no formal education are generally illiterate and more likely to be health illiterate, which subsequently lead to poorer outcomes especially in a chronic disease like AF.^11,27^

Low education and low income often coincide. Low income could lead to lack or poor choice of health insurance. In Thailand, such that consequences were broken down since the implementation of UCS. UCS provides healthcare coverage to the unemployed who primarily are in the low-income group. According to the 2004 national health insurance data, half of UCS insurers were in the bottom two poorest quintiles while half of CSMBS insurers were in the richest quintile.^28^ Unlike other insurance plans for the poor, UCS provides extensive and comprehensive benefit packages.^6^ High-cost services, such as percutaneous cardiac intervention, heart transplantation, or catheter ablation are currently covered by UCS.^2,6^ One of the remaining discrepancies between UCS and CSMBS packages is the drug plan. UCS restricts access to medications outside the National List of Essential Medicines, notably DOACs, while CSMBS covers with some conditions.^2,22^ In our AF patients, the discrepancies in the coverages did not result in outcome disparities between insurance plans. UCS insurers had essentially the same RMSTs of all-cause mortality, ischemic stroke, and major bleeding as insurers of other plans.

Outcome disparities between UCS and CSMBS have been previously reported.^7-9^ In the retrospective analysis from 2010 national insurance database, 5-year mortality after AF admission was higher in UCS than in CSMBS.^8^ Here, our patients were prospectively enrolled and structurally followed up. The enrollment took place after the 3^rd^ strategic plan of UCS began in 2012.^6^ During the plan, the benefits were expanded to seasonal influenza vaccine, screening of complications from diabetes and hypertension, long-term care for dependent elderly, and etc. The differences in trial designs and time periods of enrollment could explain the discrepancies in the results of ours and prior trials.

We reported the effect size of educational disparities in months loss. The numbers may appear small but they are time loss over a fixed period of 36 months. Losing 1.8 months in survival time for those with no formal education would therefore mean losing 5% of their time. Compared to the health production function analysis of 35 members countries of Organisation for Economic Co-operation and Development countries,^11^ a 10% increase in education of the population was associated with a gain of months in life expectancy and a 100% increase with a gain of 23.8 months.

Our results provide some insights to the national health insurance policy. We showed that in a moderate-to-high income country like Thailand, the disparity in insurance coverage could be reduced or even removed by a well maintained pro-poor health insurance like UCS. The findings also emphasize the utmost importance of education. Education does not only educate people, but also makes them healthier.^11^ For a chronic condition like AF, adherence to the therapy requires some understanding towards the disease which entails education.^14^ In our patients, the education level associated with better clinical outcomes was higher education, the level attained only in a minority of Thai population. According to the government reports,^3^ less than 25% of the working age population attained higher education and more than 20% of students left schools after completion of the compulsory lower secondary level. Clearly, there is a plenty of room for improvement to make education more inclusive and accessible.

### Strength & Limitations

Our trial is prospective and nationwide. The enrollment and follow-up were well structured and validated. For survival analysis, we chose RMST, rather than Log-rank test or Cox-proportional hazard model, to avoid the question of non-proportionality. The truncation time for the analysis was the pre-specified follow-up time and therefore optimal for the performance of the test.^20^

We studied 2 sociodemographic indicators, educational attainment and insurance plans. Both are important factors but could possibly be confounded by others such as income, living environment, and etc. Additionally, patients were enrolled through medical facilities, thus excluding those who could not or decided not to access the health care.

There were differences in many baseline comorbidities. Even after the adjustment in the multivariate analysis, some residuals confounders may remain. Due to the trial design, associations of education with clinical outcomes, rather than causes and effects, can only be assumed.

## Conclusions

In a large nationwide prospective cohort of AF patients, educational attainment was associated with all-cause mortality and ischemic stroke independent of multiple covariates. Over a period of 36 months, RMSTs to all-cause mortality and ischemic stroke of those with higher education were extended by 1.8 and 1 months respectively compared to those without education. Incidences of all-cause mortality, ischemic stroke, and major bleeding did not differ across all types of insurance plans.

## Data Availability

The individual anonymized data supporting the analyses contained in the manuscript will be made available upon reasonable written request.

## Footnotes

### Competing Interests

None

### Contributorship

SA and RK had the idea for and designed the study and were responsible for the overall content as guarantors. All authors collected the data. SA performed the statistical analysis. SA mainly wrote the manuscript with support from TT, KJ, and RK. All authors provided critical feedback and contributed to the final manuscript.

## Acknowledgement

None

## Sources of Funding

This study was funded by grants from the Health Systems Research Institute (HSRI; grant no. 59-053), the Heart Association of Thailand under the Royal Patronage of H.M. the King, and the Royal College of Physicians of Thailand.

## Patient and public involvement

Patients or the public were not involved in the design, or conduct, or reporting, or dissemination plans of our research.

## Patient consent for publication

Not required.

## Ethics approval

The protocol for this study was approved by the institutional review boards (IRBs) of the Thailand Ministry of Public Health (which covers IRBs for Buddhachinaraj Hospital, Chiangrai Prachanukroh Hospital, Chonburi Hospital, Lampang Hospital, Maharat Nakorn Ratchasima Hospital, Nakornping Hospital, Prapokklao Hospital (Chanthaburi), Ratchaburi Hospital, Surat Thani Hospital, Surin Hospital, Udonthani Hospital, and Sapphasitthiprasong Hospital) and Central Research Ethics Committee (CREC, which covers IRBs for Central Chest Institute of Thailand, Charoen Krung Pracha Rak Hospital, Chiang Mai Hospital, King Chulalongkorn Memorial Hospital, Naresuan University Hospital, Songklanakarind Hospital, Ramathibodi Hospital, Siriraj Hospital, Thammasat Hospital, Golden Jubilee Medical Center, Srinakarind Hospital, Phramongkutklao Hospital, Police General Hospital, and Faculty of Medicine Vajira Hospital) and IRB of Queen Savang Vadhana Memorial Hospital. All patients provided written informed consent prior to participation.

---

STROBE Statement–Checklist of items that should be included in reports of ***cohort studies***

|  | Item No | Recommendation | Page No |
| --- | --- | --- | --- |
| **Title and abstract** | 1 | (a) Indicate the study's design with a commonly used term in the title or the abstract<br>(b) Provide in the abstract an informative and balanced summary of what was done and what was found | 1-2 |
| **Introduction** |  |  |  |
| Background/rationale | 2 | Explain the scientific background and rationale for the investigation being reported | 4-5 |
| Objectives | 3 | State specific objectives, including any prespecified hypotheses | 4-5 |
| **Methods** |  |  |  |
| Study design | 4 | Present key elements of study design early in the paper | 5 |
| Setting | 5 | Describe the setting, locations, and relevant dates, including periods of recruitment, exposure, follow-up, and data collection | 5 |
| Participants | 6 | (a) Give the eligibility criteria, and the sources and methods of selection of participants. Describe methods of follow-up<br>(b) For matched studies, give matching criteria and number of exposed and unexposed | 5 |
| Variables | 7 | Clearly define all outcomes, exposures, predictors, potential confounders, and effect modifiers. Give diagnostic criteria, if applicable | 5-6 |
| Data sources/<br>measurement | 8* | For each variable of interest, give sources of data and details of methods of assessment (measurement). Describe comparability of assessment methods if there is more than one group | 5-6 |
| Bias | 9 | Describe any efforts to address potential sources of bias | 5-7 |
| Study size | 10 | Explain how the study size was arrived at | 5-6 |
| Quantitative variables | 11 | Explain how quantitative variables were handled in the analyses. If applicable, describe which groupings were chosen and why | 6-7 |
| Statistical methods | 12 | (a) Describe all statistical methods, including those used to control for confounding<br>(b) Describe any methods used to examine subgroups and interactions<br>(c) Explain how missing data were addressed<br>(d) If applicable, explain how loss to follow-up was addressed<br>(e) Describe any sensitivity analyses | 6-7 |
| **Results** |  |  |  |
| Participants | 13* | (a) Report numbers of individuals at each stage of study—eg numbers potentially eligible, examined for eligibility, confirmed eligible, included in the study, completing follow-up, and analysed<br>(b) Give reasons for non-participation at each stage<br>(c) Consider use of a flow diagram | 7 |
| Descriptive data | 14* | (a) Give characteristics of study participants (eg demographic, clinical, social) and information on exposures and potential confounders<br>(b) Indicate number of participants with missing data for each variable of interest<br>(c) Summarise follow-up time (eg, average and total amount) | 7-8 |
| Outcome data | 15* | Report numbers of outcome events or summary measures over time | 8-10 |
| Main results | 16 | (a) Give unadjusted estimates and, if applicable, confounder-adjusted estimates and their precision (eg, 95% confidence interval). Make clear which confounders were adjusted for and why they were included<br>(b) Report category boundaries when continuous variables were categorized<br>(c) If relevant, consider translating estimates of relative risk into absolute risk for a meaningful time period | 7-10 |
| Other analyses | 17 | Report other analyses done—eg analyses of subgroups and interactions, and sensitivity analyses | 9-10 |
| **Discussion** |  |  |  |
| Key results | 18 | Summarise key results with reference to study objectives | 10 |
| Limitations | 19 | Discuss limitations of the study, taking into account sources of potential bias or imprecision. Discuss both direction and magnitude of any potential bias | 13 |
| Interpretation | 20 | Give a cautious overall interpretation of results considering objectives, limitations, multiplicity of analyses, results from similar studies, and other relevant evidence | 10-12 |
| Generalisability | 21 | Discuss the generalisability (external validity) of the study results | 12 |
| **Other information** |  |  |  |
| Funding | 22 | Give the source of funding and the role of the funders for the present study and, if applicable, for the original study on which the present article is based | 20 |

\*Give information separately for exposed and unexposed groups.
**Note:** An Explanation and Elaboration article discusses each checklist item and gives methodological background and published examples of transparent reporting. The STROBE checklist is best used in conjunction with this article (freely available on the Web sites of PLoS Medicine at http://www.plosmedicine.org/, Annals of Internal Medicine at http://www.annals.org/, and Epidemiology at http://www.epidem.com/). Information on the STROBE Initiative is available at http://www.strobe-statement.org.

## Notes

### Competing Interest Statement

The authors have declared no competing interest.

### Clinical Trial

Thai Clinical Trial Registration; Study ID: TCTR20160113002

### Clinical Protocols

http://www.thaiclinicaltrials.org/

### Author Declarations

Statement of IRB approval The protocol for this study was approved by the institutional review boards (IRBs) of the Thailand Ministry of Public Health (which covers IRBs for Buddhachinaraj Hospital, Chiangrai Prachanukroh Hospital, Chonburi Hospital, Lampang Hospital, Maharat Nakorn Ratchasima Hospital, Nakornping Hospital, Prapokklao Hospital (Chanthaburi), Ratchaburi Hospital, Surat Thani Hospital, Surin Hospital, Udonthani Hospital, and Sapphasitthiprasong Hospital) and Central Research Ethics Committee (CREC)(which covers IRBs for Central Chest Institute of Thailand, Charoen Krung Pracha Rak Hospital, Chiang Mai Hospital, King Chulalongkorn Memorial Hospital, Naresuan University Hospital, Songklanakarind Hospital, Ramathibodi Hospital, Siriraj Hospital, Thammasat Hospital, Golden Jubilee Medical Center, Srinakarind Hospital, Phramongkutklao Hospital, Police General Hospital, and Faculty of Medicine Vajira Hospital) and IRB of Queen Savang Vadhana Memorial Hospital. All patients provided written informed consent prior to participation.

